# Dance Interventions for Individuals Post-Stroke - A Scoping Review Protocol

**DOI:** 10.1101/2021.05.26.21257850

**Authors:** Danielle Kipnis, Helena Kruusamäe, Miriam King, Abigail Schreier, Lori Quinn, Hai-Jung Steffi Shih

## Abstract

The purpose of this scoping review will be to explore the current literature on dance interventions for individuals post-stroke. Specifically, we will conduct a systematic search of published studies to map the state of the literature on feasibility, intervention procedures, and efficacy of dance to improve health-related outcomes for individuals post-stroke. Stroke is the leading cause of long-term disability frequently causing motor and cognitive impairments that impact functional abilities. Dance inherently encompasses key principles of motor learning including cognitive-motor specific task practice, sensory feedback, and social engagement. Dance and music create an enriched environment, engaging emotion and promoting positive affect.

Electronic databases were searched in February 2021. Original, peer-reviewed studies will be included if they broadly describe the use of a dance intervention for individuals post-stroke. Studies including other neurological populations will be considered only if stroke results can be isolated. We will categorize studies based on dance style, study type, population characteristics, and study setting. We will narratively synthesize results, assessing feasibility, intervention procedures, and efficacy of dance to improve health-related outcomes in stroke survivors, including potential motor, cognitive, psychological, and social benefits.

This scoping review will be the first to broadly describe the existing literature on dance interventions for individuals post-stroke. We hope to identify trends in outcomes measures as well as reveal successes, limitations, and gaps in the literature to inform potential directions for future research in the field.

## Introduction

Stroke is the leading cause of long-term disability (Simpson et al., 2011). Stroke can affect multiple brain regions and lead to severe motor impairments, which greatly impact daily function and quality of life (Campbell & Khatri, 2020). Reduced participation in the community after stroke also leads to social isolation, depression, lack of confidence, and decreased autonomy (Carod-Artal et al., 2000; Salter et al., 2008). Furthermore, sedentary lifestyles due to motor impairments can increase the risk of recurrent strokes and cardiovascular disease, two leading causes of mortality among individuals post-stroke (Simpson et al., 2011). Traditional rehabilitation strategies used in physical and occupational therapy can effectively improve functional outcomes in individuals post-stroke (Cickusic et al., 2015; Cook et al., 2005). However, low patient adherence and sustainability remain limitations for these rehabilitative modes due to limited community-based options, patient motivation, and social engagement (Simpson et al., 2011). Therefore, development and testing of exercise-based interventions that are accessible, promote mobility and enjoyment are warranted to improve and sustain health-related outcomes in individuals post-stroke.

Dance as a multi-modal intervention has promising physical, psychological, cognitive, and social benefits that can lead to neuroplastic changes (Teixeira-Machado et al., 2019). A review by Keogh et al. suggests that dance can significantly improve aerobic power, muscle endurance, strength, lower body flexibility, gait speed, and static and dynamic balance for older adults (Keogh et al., 2009a). Individuals who engage in dancing into older age better preserve cognitive and perceptual abilities than those who do not dance or participate in sports (Kattenstroth, 2010). Perhaps most importantly, dance encourages self-expression, promotes social interaction, and facilitates a sense of community. This combination of physical activity with emotional expression, social interaction, motor skill acquisition, and music creates an enriched environment that may promote neuroplastic changes in the injured brain (Keogh et al., 2009b). In animal studies, exercise in combination with an enriched environment has an additive effect on the brain: physical activity alone can only stimulate precursor cells for adult neurogenesis, whereas the addition of an enriched environment promotes survival of these immature cells (Kempermann et al., 2010).

Research on dance for people with Parkinson’s (PwP) and other neurological conditions (Patterson et al., 2018a) suggest the effects of dance for healthy older adults can be generalized to neurological populations. A meta-analysis by Shanahan et al. on dance for PwP favors dance over control interventions to improve balance (Shanahan et al., 2015). Additionally, dance interventions for PwP have resulted in greater improvements in balance than healthy older adults (Earhart, 2009), indicating that dance could have more profound benefits in neurological populations. Dance has similarly improved balance for individuals with spinal cord injury (Sapezinskiene et al., 2009), spatiotemporal gait parameters and motor function for individuals with Huntington’s disease (Trinkler et al., 2019) and mobility for individuals with multiple sclerosis (Mandelbaum et al., 2016). Finally, studies on dance for PwP and dementia suggest it enhances social networks, contributes to improved quality of life, and could improve motivation to continue exercise-related activities (Earhart, 2009; Klimova et al., 2017; Shanahan et al., 2015).

Despite evidence in other neurological populations, dance is under-researched as a rehabilitation strategy for individuals post-stroke. This is possibly because individuals post-stroke often have sudden and severe motor impairment rather than progressive degeneration as in Parkinson’s disease, multiple sclerosis, Huntington’s disease, and dementia. Some preliminary work has assessed the feasibility and potential benefits of dance for individuals post-stroke. Several forms of dance have been explored, and many classes are adapted to meet post-stroke individuals’ abilities. Due to this growing body of evidence, this scoping review will evaluate and summarize the existing literature investigating dance for stroke. We believe a scoping review, rather than a systematic review, is most appropriate because research in the field of dance for stroke survivors is nascent. Particularly, publications are scarce, study design is heterogeneous, and sample sizes are small. A scoping review provides a clear framework to map the broad scope of current literature and the opportunity to include a variety of study designs while maintaining methodological rigor. In 2018, Patterson et al. summarized the evidence for dance in neurological populations other than PD (Patterson et al., 2018b). However, this review focused on studies assessing gait and balance impairments and included only three studies specific to stroke; several additional studies on stroke have since been published.

The purpose of this scoping review will be to explore the current literature on dance interventions for individuals post-stroke. Specifically, we will conduct a search of published studies to map the state of the literature on feasibility, intervention procedures, and efficacy of dance to improve health-related outcomes for individuals post-stroke. This review will identify gaps in the literature to provide recommendations for future research.

## Methods

We will follow the Arksey and O’Malley original framework for scoping reviews and present the five recommended steps in the following sections (Arksey & O’Malley, 2005). Additionally, we will adhere to the Preferred Reporting Items for Systematic Reviews and Meta-Analyses Extension for Scoping Reviews (PRISMA-ScR) checklist when reporting our findings (Tricco et al., 2018).

### Identify the Research Question

Research indicates dance may be an effective intervention for neurological and healthy populations (Aguiar et al., 2016; Bruyneel, 2019; Rehfeld et al., 2018), with growing evidence on the effects of dance for people post-stroke. Therefore, a scoping review is necessary to map the results from emerging evidence, identify gaps, and suggest future directions for the field. To achieve this goal, we will pose the following research question: “What is known from the existing literature about the feasibility, intervention procedures, and efficacy of dance interventions to improve health-related outcomes for individuals post-stroke?” Health-related outcomes may include, but are not limited to, motor, cognitive, psychological, and social benefits.

### Identify Relevant Studies

The eligibility criteria used in this review are as follows: (1) studies broadly describe the use of a dance intervention for individuals post-stroke; (2) studies can include any health-related outcome; (3) studies must be written in English; (4) studies can include other populations, however they are ineligible if the results for adults post-stroke cannot be isolated; and (5) studies must be original research. If a relevant systematic review (such as dance for neurological population) is found, we will only include original studies within the review that meet eligibility criteria. For the purpose of this review, we will include studies where researchers define the intervention as dance. We have specifically excluded other movement modalities such as yoga, Pilates, and Tai Chi. Though these modalities are similar to dance – they incorporate whole body movements and can sometimes be rhythmic and use music – they are defined styles that are different from dance.

The following search strategy will be used to search for relevant studies that include keywords in the title, abstract, or full-text: dance OR “dance movement therapy” AND stroke OR cerebrovascular disorder/accident. The databases selected for this review are Pubmed, Scopus, Google Scholar, Proquest, MedRxiv and CINHAL (see Supplementary Table for individual search strategies). Since Google Scholar is not a refined index, we will include the first five pages of the search listed by relevance. We chose these databases to include peer-reviewed articles, dissertations, conference proceedings, and pre-print manuscripts to broaden the scope of this search.

### Study selection

Studies identified through database search will be imported into the reviewer tool Covidence v.2603 (Covidence.org, Melborne, Australia) and duplicates will be automatically removed. Covidence is an online tool that helps to sort and share studies to facilitate the screening process. Pairs of independent reviewers will first screen the titles and abstract from a list of articles obtained by using our key search terms. The remaining studies will then be full-text screened by pairs of reviewers. Finally, we will screen reference lists of included studies for additional relevant articles not found in the initial search and possible inclusion in the review. Any conflicts or discrepancies throughout both rounds will be discussed and resolved by the group. A PRISMA flow diagram will be used to report the study selection process (Tricco et al., 2018).

### Data charting

Relevant data will be extracted using customized templates in a spreadsheet format. Each article will be read, reviewed, and charted by two independent reviewers. Table 1 summarizes the specific data and details included on the data charting spreadsheet. We will not perform a quality assessment of included studies because it is not required for scoping reviews and we expect included studies will too heterogeneous to do so.

**Table 1.** Data to be extracted.

| Data | Details to be extracted (if available) |
| --- | --- |
| Publication Summary | Author, title, year of publication, journal, eligibility criteria, type of research (e.g., intervention, qualitative, review, case-study dissertation) |
| Population | Total sample size, age, gender, race, ethnicity, medication, diagnosis, time since diagnosis and/or cerebrovascular event, caregiver present (yes/no), mobility device |
| Intervention | Study aims or purpose, intervention and comparator, frequency, duration, intensity, methodology, instructor characteristics (e.g., dance training, experience with stroke), dance style or genre, use of music, individual, group, partner, non-partnered |
| Setting | Location (e.g., healthcare facility, community center, dance studio, home), method of delivery (e.g., virtual, home-based, exergaming, in-person), cost and equipment, accessibility |
| Outcomes | Measures and results: physical function, psychosocial, cognitive, quality of life, relationships (e.g., family, caregiver); statistical analysis, limitations or barriers, adverse events |
| Bias | Each source individually reviewed for potential source of bias and/or methodological weakness |
| Other | Adherence, feasibility, clinical applicability |

### Collating, summarizing, and reporting the results

We will narratively summarize the data as well as provide tables and figures. The extracted data will also be visualized using bubble plots, pie charts, tables, and graphs where appropriate. Concept and themes will be determined through an iterative process. Terms will be defined where needed. Studies will be categorized based on dance intervention style (e.g. dance rehabilitation therapy, dance-based exergaming), study type (e.g. qualitative, quantitative, case description), population characteristics (e.g. acute or chronic stroke) and/or study setting (e.g. in-patient, out-patient, community-based).We will discuss the scope of the current literature and strengths and weaknesses of data extracted to identify gaps in the field of dance for stroke and make recommendations for future research. These steps follow recommendations for scoping reviews by Levac et al., (Levac et al., 2010).

## Results

Database search based on title, abstract, keywords, and body text was performed on February 15th and 18th, 2021. Title, abstracts, and full-texts have been screened by at least two reviewers. Data extraction and data charting has begun. Completion is expected in Fall 2021.

## Discussion

Evidence supports the importance of aerobic and rehabilitative exercise for individuals post-stroke as a means to promote motor learning, community re-integration, and subsequent stroke prevention (Eng et al., 2008; Saunders et al., 2013). There is a substantial body of evidence demonstrating that dance is uniquely multi-modal and may have additional health-related benefits compared to traditional exercise in healthy older adults and several neurological populations such as Parkinson’s disease, multiple sclerosis, Huntington’s disease, and dementia. Dance inherently encompasses key principles of motor learning including cognitive-motor specific task practice, sensory feedback, and social engagement (Lewthwaite, 2010; Magill & Anderson, 2016). Despite this evidence, our understanding of the effects of dance for individuals post-stroke is limited.

Evidence in this field has begun to grow in the last several years. This scoping review will be the first to broadly describe the existing literature on dance interventions for individuals post-stroke. We anticipate describing key components of dance interventions, including intervention timing, dance style, and setting. Additionally, we hope to identify trends in outcomes measures related to potential motor, cognitive, psychological, and social benefits of dance. Finally, in synthesizing this data more broadly, we aim to reveal successes, limitations, and gaps in the literature to inform potential directions for future research in the field.

## Data Availability

All extracted/charted data are available upon request.

## Notes

**Conflict of interest:** The authors declare no conflict of interest.

### Competing Interest Statement

The authors have declared no competing interest.

### Clinical Trial

N/A

### Funding Statement

No external funding was received.

### Author Declarations

This scoping review is exempt from IRB approval.

