## Supplementary Table for "Dance Interventions for Individuals Post-Stroke - A Scoping Review Protocol"

**Supplementary Table. Search terms by Database**

| Database | **Search Terms** |
| --- | --- |
| Pubmed | (dance OR “dance movement therapy”) AND (stroke OR “cerebrovascular accident”) |
| Scopus | (dance OR “dance movement therapy”) AND (stroke OR “cerebrovascular accident”) |
| Google Scholar* | (dance AND stroke AND (therap* OR intervention OR rehab*) |
| Proquest | (noft(dance) OR noft(dance movement therapy)) AND (noft(stroke) OR (cerebrovascular)) |
| MedRxiv | (dance AND stroke) |
| CINHAL | (dance OR “dance movement therapy”) AND (stroke OR “cerebrovascular accident”) |

**Include first 5 pages sorted by relevance*
